# Estimating the epidemiology of chronic Hepatitis B Virus (HBV) infection in the UK: what do we know and what are we missing?

**DOI:** 10.1101/2022.04.12.22273568

**Authors:** Cori Campbell, Tingyan Wang, Rebekah Burrow, Sema Mandal, Julia Hippisley-Cox, Eleanor Barnes, Philippa C Matthews

**Affiliations:** Nuffield Department of Medicine, University of Oxford, Oxford, UK; NIHR Oxford Biomedical Research Centre, University of Oxford, Oxford, UK; QResearch / Nuffield Dept of Primary Care, Oxford, UK; Blood safety, Hepatitis, STI & HIV Division, UK Health Security Agency; National Institute for Health Research Health Protection Unit, London, UK; Department of Hepatology, Oxford University Hospitals, John Radcliffe Hospital, Headley Way, Headington, Oxford OX3 9DU; The Francis Crick Institute, 1 Midland Road, London NW1 1AT, UK; Division of Infection and Immunity, University College London, Gower Street, London, WC1E 6BT, UK; Department of Infectious Diseases, University College London Hospital, Euston Road, London NW1 2BU, UK

**Keywords:** hepatitis B virus, HBV, prevalence, epidemiology

## Abstract

**Background:** HBV is the leading global cause of cirrhosis and primary liver cancer. The virus’s attributable disease burden in the UK is concentrated in vulnerable populatons including ethinic minorities, people experiencing homelessness and people born in high-prevalence countries. Despite this the UK HBV population has not been well characterised, and estimates of UK HBV prevalence and/or positivity rate vary widely across sources. We summarised datasets that are available to represent UK CHB epidemiology, consider differences between sources, and discuss deficiencies in current estimates.

**Methods:** We searched for estimates of CHB case numbers in the UK (incorporating incidence and/or prevalence-like data) across a range of available sources, including UK-wide reports from government bodies, publications from independent bodies (including medical charities and non-governmental organisations) and articles in peer-reviewed scientific journals. We present positivity rates from each respective data source but caution that estimates may not be representative of the true UK-wide population prevalence.

**Results and Discussion:** Six CHB case number estimates were identified, with three estimates reporting information concerning population subgroups, including number of infected individuals across age, sex and ethnicity categories., Estimates among sources reporting prevalence varied from 0.27% to 0.73%. An alterantive proxy for population prevalence (obtained via the UK antenatal screening programme which achieves over 95% coverage of every pregnant woman) estimated a CHB prevalence of <0.5%. Estimates varied by sources of error, bias and missingness, data linkage, and substantial “blind spots” in consistent testing and registration of HBV diagnoses. Multi-parameter evidence synthesis and back-calculation model methods similar to those used to generate estimates of HCV ad HIV population-wide prevalence may be applicable to HBV.

## Background

Hepatitis B virus (HBV) is the leading global cause of cirrhosis, and of primary liver cancer incidence and mortality (1,2). Nearly 300 million individuals worldwide are estimated to be living with chronic HBV (CHB) infection. Risks of complications and death are mitigated by screening to detect cases of infection, clinical monitoring of chronic infection (including liver cancer surveillance in high-risk cases), and antiviral therapy in those who meet treatment criteria (3).

The United Kingdom (UK) is regarded as a low-prevalence setting for CHB (3). However, the attributable disease burden may be substantial in specific population subgroups including people who inject drugs, the prison population, people experiencing homelessness, and individuals belonging to minority ethnic groups and born in countries where the prevalence of CHB is higher (4). Thus, CHB is concentrated in potentially vulnerable and/or disadvantaged population subgroups.

Epidemiological characterisation of the UK CHB population has been limited, with no central registry of infected persons. Existing data may primarily reflect new diagnoses (a combination of incident acute infection and new diagnoses of chronic infection), but caution is needed in making inferences about prevalence. Accurate estimation of prevalence is challenging, because complete HBV data are not likely to be well captured by large-scale electronic health record (EHR) databases for either primary or secondary care (5), as many CHB cases remain untested and therefore undiagnosed.

The World Health Organization (WHO) has set targets for viral hepatitis elimination within its Sustainable Development Goals for 2030. The Global Health Sector Strategy on Viral Hepatitis (6) identifies specific goals, including diagnosis in 90% of chronic infections, 90% reduction in incidence of chronic infection, and 80% treatment coverage in those eligible. High quality epidemiological data are therefore crucial to focus and measure progress, inform policy and interventions, reduce inequities and underpin resource allocation. We herein summarise datasets that are available to represent UK CHB epidemiology, consider differences between sources, and discuss deficiencies in current estimates.

## Methods

We searched for estimates of CHB case numbers in the UK (incorporating incidence and/or prevalence-like data) across a range of available sources. We included UK-wide reports from government bodies, publications from independent bodies (including medical charities and non-governmental organisations) and articles in peer-reviewed scientific journals. We present positivity rates from each respective data source, but caution that these estimates are not representative of the true UK-wide population prevalence. Details of study samples/denominator are provided. The Office for National Statistics (ONS) provides UK population estimates as a point of reference for the overall denominator (7).

We also utilised data from the UK primary care database QResearch, which contains over 35 million patient records from more than 1800 individual practices (7). QResearch was established in 2002 and contains anonymised individual-level patient EHR. Data are collected prospectively and are linked to hospital episode statistics (HES), National Cancer Registration Analysis Service (NCRAS) and ONS mortality data. QResearch ethics approval is with East Midlands-Derby Research Ethics Committee (reference 18/EM/0400).’

We identified individuals in the QResearch (version 44) database who had a record of a diagnostic Systemised Nomenclature of Medicine (SNOMED)/Read or International Classification of Disease (ICD) code indicative of CHB, or who had a history of ≥1 hepatitis B surface antigen (HBsAg) or viral load (VL) measurement. From this sample we identified individuals between 01 January 1999 and 31 December 2019, age ≥18 years with CHB, defined as: i) record of a diagnostic SNOMED/Read code indicating CHB; or ii) record of a diagnostic ICD-9 or −10 code indicating CHB; and/or iii) Presence of HBsAg or VL on ≥2 recordings ≥6 months apart. The characteristics of HBV infection in the cases we identified are further described elsewhere (7).

We have also drawn on findings from a similar investigation previously undertaken in the Clinical Practice Research Datalink (CPRD) (8), which is another UK primary care database containing EHRs for over 16 million patients. This previous investigation identified CHB individuals from patients registered in the database between 2000 and 2015.

## Results and discussion

UK data for CHB epidemiology are summarised in Table 1. Three of six estimates report information concerning population demographics, including number of infected individuals across age, sex and ethnicity categories. Among sources setting out to report prevalence, estimates varied from 0.27% (British Liver Trust / Department of Health and Social Care (DHSC) 2002 estimate) to 0.73% (estimate by the Polaris Institute). An alterantive proxy for population prevalence is obtained via the UK antenatal screening programme, which achieves over 95% coverage of every pregnant woman annually (approx. 700,000 women in the UK), with a CHB prevalence of <0.5% (9). Differences between sources highlights varied sources of error, bias and missingness, problems with data linkage, and substantial “blind spots” in consistent testing and registration of HBV diagnoses.

**Table 1.**
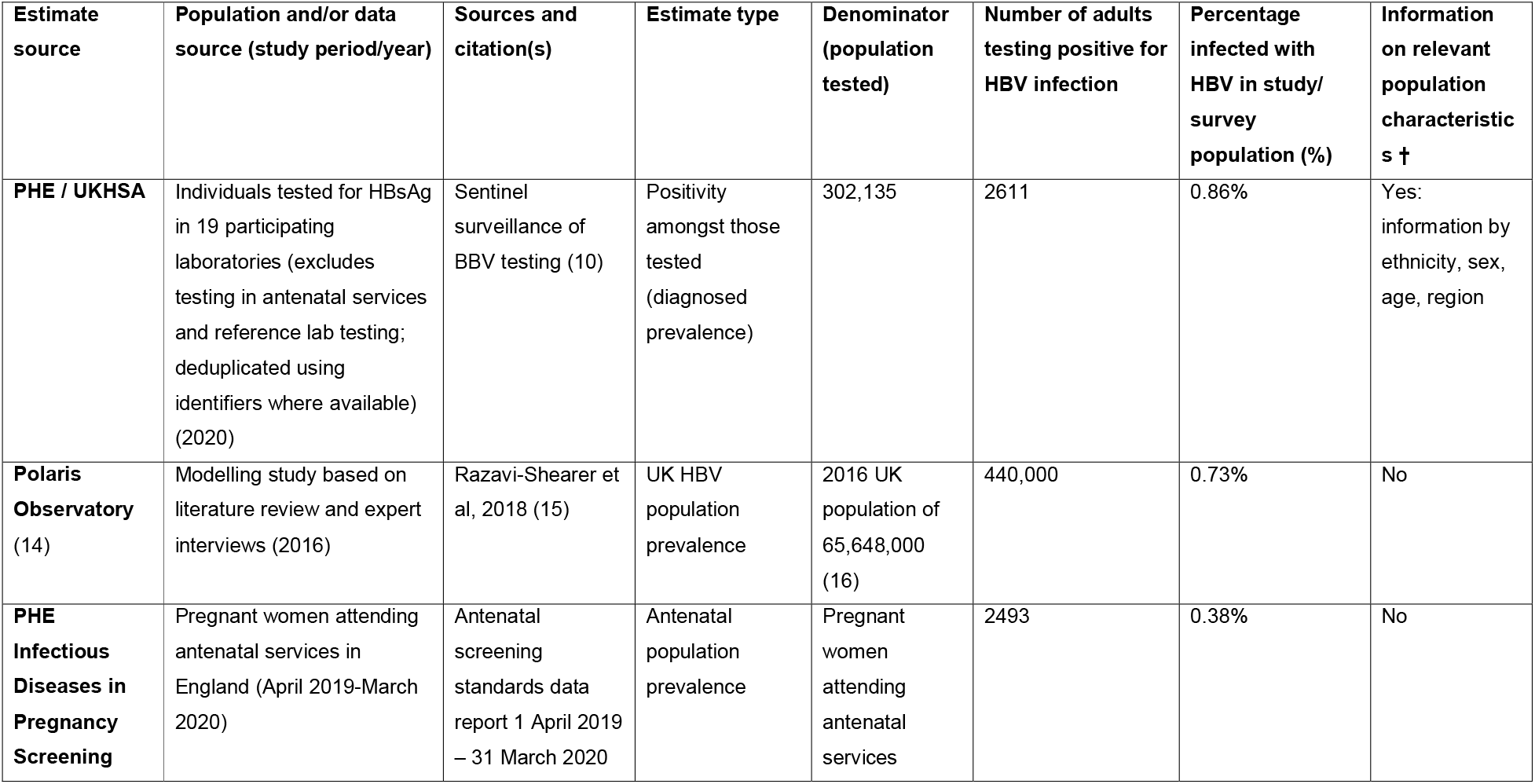
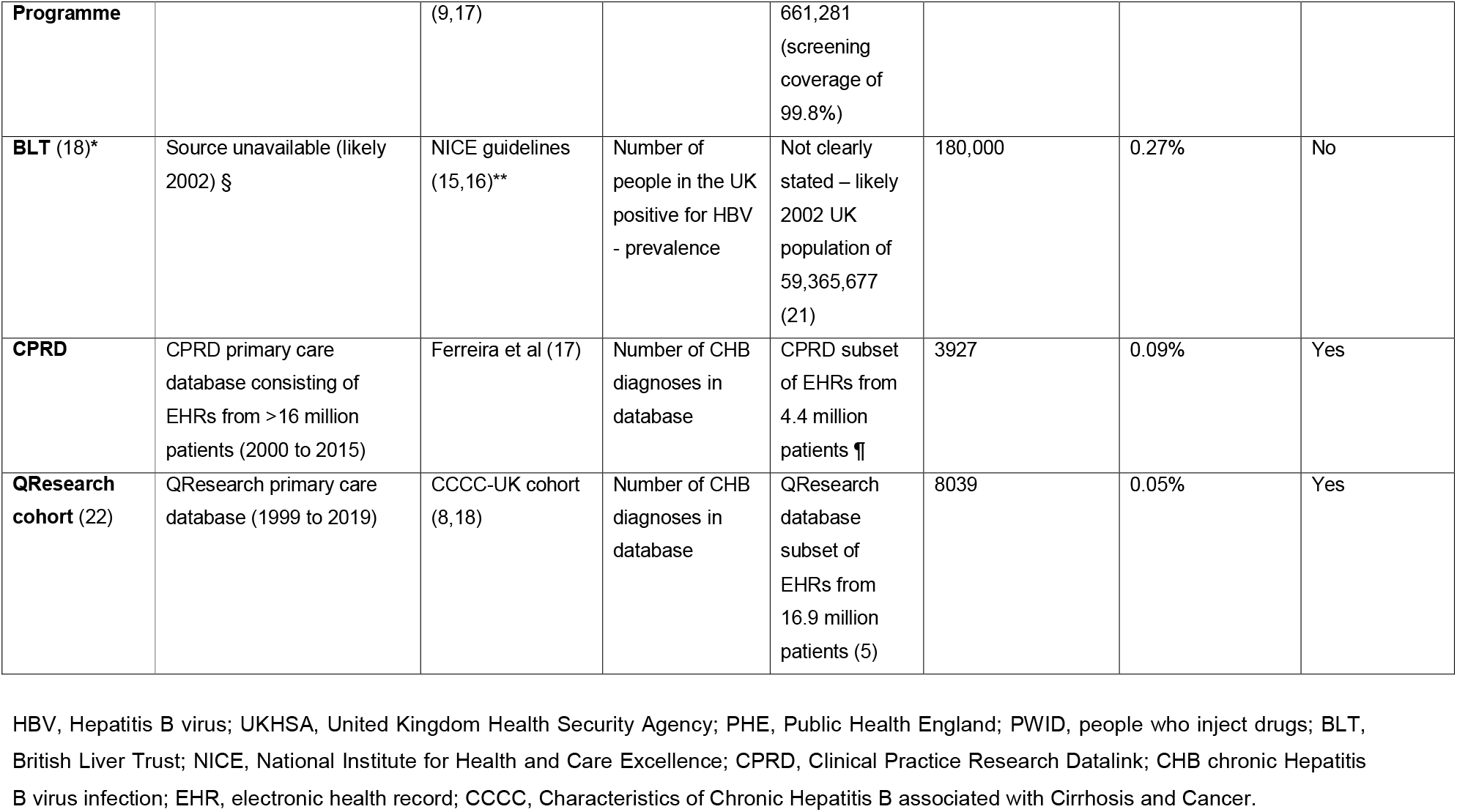
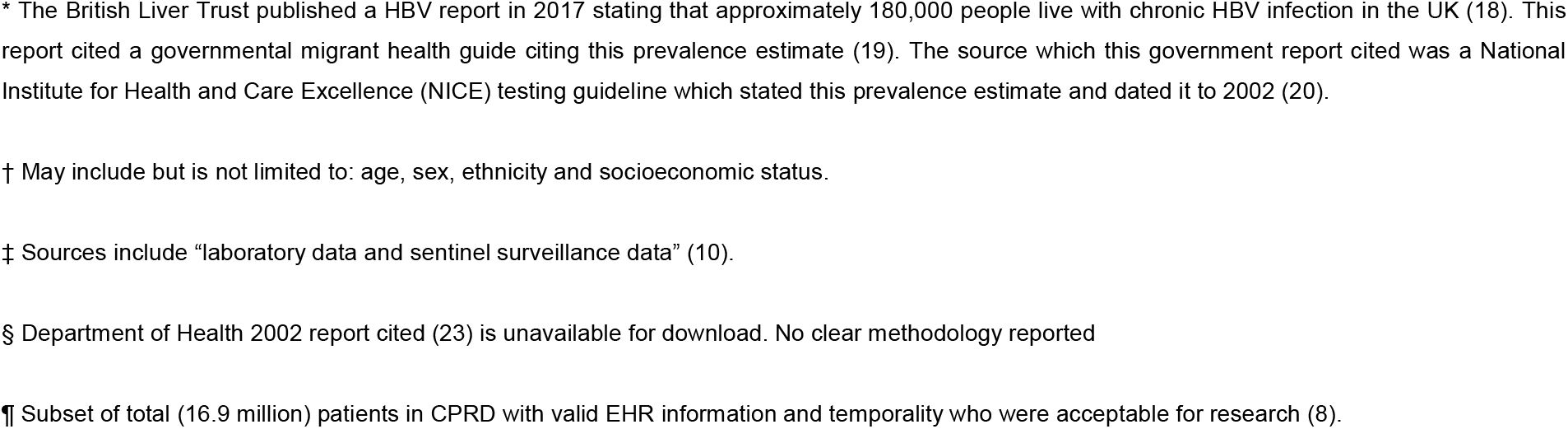
Prevalence estimates for chronic hepatitis B virus (HBV) infection in sub-populations in the United Kingdom.

As HBV is a notifiable disease in the UK, the UK Health Security Agency, UKHSA (previously Public Health England, PHE), has a comprehensive surveillance system for monitoring burden of CHB, by monitoring testing, blood donor screening and diagnoses across the care pathway. This incorporates data from diagnoses through to outcomes, (including end-stage liver disease, transplantation, liver cancer and deaths) using laboratory testing surveillance, hospital activity datasets and registries (sentinel surveillance of blood-borne virus (BBV) testing, new laboratory diagnoses, hospital episode statistics, ONS cancer and deaths registries). However, these data have not yet been combined and incorporated in a statistical model to estimate prevalence. Sentinel surveillance captures testing in community, primary care and secondary care settings across a network of laboratories covering approximately 40% of the general population of England (10). This likely gives the best estimate of diagnosed prevalence among a tested population, but because it combines acute incident infections and new diagnoses of pre-existing chronic infection, incidence and prevalence cannot be disaggregated.

The majority of diagnostic data are generated through testing individuals with risk factors for HBV infection or evidence of liver disease, and are therefore at risk of over-estimating true prevalence. However, no existing estimates factor in the undiagnosed burden, which represents the majority of people living with HBV infection (the WHO estimates that only 10.5% of people with CHB are aware of their infection status (3)). Furthermore, the highest prevalence of CHB is in groups for whom provision of healthcare is inadequate, and/or access to healthcare is challenging (including migrants, sex-workers, prisoners, and people experiencing homelessness), so overall there are still many gaps in the data, and it is most likely that estimates using primary care datasets considerably underestimate the true burden. In contrast, prevalence or test positivity among those accepting risk-based testing (as captured in laboratory testing surveillance) likely overestimates the overall population prevalence.

While UKHSA surveillance data may include some demographic characteristics (age, sex, postcode for deprivation), unless linked to other healthcare datasets, they typically lack more detailed clinical and demographic indicators (for example, measures of deprivation, lifestyle factors, assessment of liver disease, and HBV treatment coverage) which are needed to characterise the infected population. In constrast, EHR databases (such as CPRD and QResearch) have the advantage of collecting relevant demographic and clinical metadata which are not captured by UKHSA. However, linkage across data sources is disaggregated, and thereby each EHR-based estimate misses a portion of the infected population. For example, primary care data may not reflect testing conducted in secondary care (11), blood safety (transfusion/transplantation) and laboratory data generated by other services, while secondary care data are typically only reliable for the sub-population enrolled in consistent hospital follow-up. Poor data flow between diagnostic testing and EHR reflect a low clinical follow-up rate following a positive HBsAg test. This limited linkage to care reflects how services may not provide well for the CHB population, with gaps in referral pathways, inadequate communication and education (including translation services), and failures to deliver services to marginalised communities. Therefore, EHR databases offer the potential to characterise a subset of those infected with HBV, but do not currently generate a picture that is generalisable to the wider infected population, and cannot on their own be used to estimate prevalence.

Prevalence estimates for Hepatitis C virus (HCV) (12) and human immunodeficiency virus (HIV) (13) have recently been generated using multi-parameter evidence synthesis and back-calculation models. Similar modelling approaches to produce estimates of HBV incidence and prevalence in the UK are warranted. Enhanced investment is needed to support the establishment of national registries with robust centralised data linkage between sources including national laboratory surveillance systems of BBV testing and new diagnoses, and thus determine which population subgroups are bearing the majority of the HBV disease burden. This will inform prevalence modelling and provide an evidence base for delivery of appropriate resources and interventions, and to benchmark progress towards elimination targets.

**Summary box**

**Recommendations for the generation of enhanced insights into national CHB caseload**

- Expansion of systematic screening, including opportunistic approaches (sexual health, antenatal, emergency medicine, people born in high-prevalence settings).
- Improved centralised data linkage between services, including laboratory records, blood and transplant services, primary and secondary care, supported by collection of metadata.
- Disaggregation of incidence/prevalence data where possible at source.
- Establishment of regional and/or national registries to collate linked data for HBV infection at a population level and within high risk groups.
- Mathematical modelling to optimise use of existing data to generate incidence and prevalence estimates, identify systematic data gaps, refine allocation of resources and predict progress towards elimination targets.

## Data Availability

Only CC, TW, RB and JH-C have access to the QResearch individual-level patient data in order to ensure confidentiality of personal and health information, in accordance with the relevant licence agreements. QReseearch data access is according to the information on the QResearch website (www.qresearch.org).

## Ethics approval

QResearch ethics approval is with East Midlands-Derby Research Ethics Committee (reference 18/EM/0400).

## Author contrubutions

PM and CC conceptualised the study. CC conducted the literature search for estimates, and drafted the manuscript with PM. JH-C, RB. SM, TW and EB provided methodological input. JH-C, SM, TW and EB revised the manuscript.

## Acknowledgements

Access to the data was facilitated by the PHE Office for Data Release. We acknowledge the contribution of EMIS practices who contribute to QResearch® and EMIS Health and the CHANCELLOR MASTERS AND SCHOLARS OF THE UNIVERSITY OF OXFORD for expertise in establishing, developing and supporting the QResearch database. United Kingdom Health Security Agency bear no responsibility for the analysis or interpretation of the data. We would like to thank Iain Gillespie at GSK for his helpful comments on the manuscript.

## References

1. Fitzmaurice C, Akinyemiju T, Abera S, Ahmed M, Alam N, Alemayohu MA, et al. The burden of primary liver cancer and underlying etiologies from 1990 to 2015 at the global, regional, and national level results from the global burden of disease study 2015. JAMA Oncology. 2017 Dec 1;3(12):1683–91.

2. Sepanlou SG, Safiri S, Bisignano C, Ikuta KS, Merat S, Saberifiroozi M, et al. The global, regional, and national burden of cirrhosis by cause in 195 countries and territories, 1990–2017: a systematic analysis for the Global Burden of Disease Study 2017. The Lancet Gastroenterology and Hepatology [Internet]. 2020 Mar 1 [cited 2021 Apr 10];5(3):245–66. Available from: www.thelancet.com/gastrohep

3. WHO. Hepatitis B Fact Sheet. [Internet]. 2021. Available from: https://www.who.int/en/news-room/fact-sheets/detail/hepatitis-b

4. Hepatitis B in London: 2016 data [Internet]. 2019 [cited 2020 Oct 2]. Available from: https://assets.publishing.service.gov.uk/government/uploads/system/uploads/attachment_data/file/801174/London_hepatitis_B_report_2016.pdf

5. Data - QResearch [Internet]. [cited 2022 Feb 6]. Available from: https://www.qresearch.org/data/

6. Global Health Sector Strategy on viral hepatitis 2016–2021. [Internet]. Geneva; 2016 [cited 2021 Jun 14]. Available from: https://apps.who.int/iris/bitstream/handle/10665/246177/WHO-HIV-2016.06-eng.pdf?sequence=1&isAllowed=y

7. Campbell C, Wang T, Barnes E, Matthews PC, Campbell C, Wang T, et al. Baseline characteristics of a large multi-site chronic HBV electronic health record-based cohort in the UK. F1000Res. 2021 Jul 26;10.

8. Ferreira G, Stuurman AL, Horsmans Y, Cattaert T, Verstraeten T, Feng Y, et al. Hepatitis B virus infection and the risk of liver disease progression in type 2 diabetic patients with potential nonalcoholic fatty liver disease: a retrospective, observational, cohort study in the United Kingdom Clinical Practice Research Datalink. Eur J Gastroenterol Hepatol. 2020 Jan 1;32(1):101–9.

9. Mandal S, Hayden I, Neal J, Cottrell S, Regan J, de Souza S, et al. Demonstrating control of perinatal transmission of hepatitis B in the UK: a low prevalence country with universal antenatal screening and selective neonatal immunisation programmes. EASL Viral Hepatitis Elimination Conference 24-25 Poster Abstract PO 75 [Internet]. 2022 Feb [cited 2022 Mar 29]; Available from: https://easl.eu/wp-content/uploads/2022/02/Viral-Hepatitis-Elimination-2022-Abstract-book.pdf

10. Public Health England. Annual report from the sentinel surveillance of blood borne virus testing in England 2020: main report. 2021 Jul 28;15(13).

11. Wang T, Smith DA, Campbell C, Freeman O, Moysova Z, Noble T, et al. Cohort Profile: National Institute for Health Research Health Informatics Collaborative: Hepatitis B Virus (NIHR HIC HBV) Research Dataset. medRxiv [Internet]. 2021 Oct 25 [cited 2022 Apr 4];2021.10.21.21265205. Available from: https://www.medrxiv.org/content/10.1101/2021.10.21.21265205v1

12. Harris RJ, Harris HE, Mandal S, Ramsay M, Vickerman P, Hickman M, et al. Monitoring the hepatitis C epidemic in England and evaluating intervention scaleLup using routinely collected data. Journal of Viral Hepatitis [Internet]. 2019 May 1 [cited 2022 Mar 16];26(5):541. Available from: /pmc/articles/PMC6518935/

13. Presanis AM, Kirwan PD, Jackson CH, de Angelis D, Health England p, D Kirwan UP, et al. Trends in undiagnosed HIV prevalence in England and implications for eliminating HIV transmission by 2030: an evidence synthesis model. The Lancet Public Health [Internet]. 2021 Oct 1 [cited 2022 Mar 16];6(10):e739–51. Available from: www.thelancet.com/

14. Hepatitis B Dashboard United Kingdom [Internet]. Center for Disease Analysis Foundation. 2021 [cited 2022 Feb 8]. Available from: https://cdafound.org/polaris-countries-dashboard/

15. Razavi-Shearer D, Gamkrelidze I, Nguyen MH, Chen DS, van Damme P, Abbas Z, et al. Global prevalence, treatment, and prevention of hepatitis B virus infection in 2016: a modelling study. Lancet Gastroenterol Hepatol [Internet]. 2018 Jun 1 [cited 2022 Feb 8];3(6):383–403. Available from: https://pubmed.ncbi.nlm.nih.gov/29599078/

16. Office for National Statistics. Population estimates for the UK, England and Wales, Scotland and Northern Ireland: mid-2016 [Internet]. 2017 [cited 2022 Mar 29]. Available from: https://www.ons.gov.uk/peoplepopulationandcommunity/populationandmigration/populationestimates/bulletins/annualmidyearpopulationestimates/mid2016

17. Public Health England. Antenatal screening standards: data report 1 April 2019 to 31 March 2020 - GOV.UK [Internet]. 2021 [cited 2022 Mar 29]. Available from: https://www.gov.uk/government/statistics/antenatal-screening-standards-data-report-2019-to-2020/antenatal-screening-standards-data-report-1-april-2019-to-31-march-2020

18. British Liver Trust. Hepatitis B. 2017.

19. Hepatitis B: migrant health guide - GOV.UK [Internet]. [cited 2022 Feb 8]. Available from: https://www.gov.uk/guidance/hepatitis-b-migrant-health-guide

20. Hepatitis B and C testing: people at risk of infection (PH43) [Internet]. 2012 [cited 2022 Feb 8]. Available from: www.nice.org.uk/guidance/ph43

21. Office for National Statistics. UK Population Estimates 1851 to 2014 [Internet]. 2015 [cited 2022 Mar 29]. Available from: https://www.ons.gov.uk/peoplepopulationandcommunity/populationandmigration/populationestimates/adhocs/004356ukpopulationestimates1851to2014

22. Characteristics of Chronic Hepatitis B associated with Cirrhosis and Cancer [Internet]. [cited 2020 Aug 18]. Available from: https://www.qresearch.org/research/approved-research-programs-and-projects/characteristics-of-chronic-hepatitis-b-associated-with-cirrhosis-and-cancer/

23. [ARCHIVED CONTENT] Getting ahead of the curve: a strategy for combating infectious diseases (including other aspects of health protection)□: Department of Health -Publications [Internet]. [cited 2022 Feb 15]. Available from: https://webarchive.nationalarchives.gov.uk/ukgwa/+/www.dh.gov.uk/en/Publicationsandstatistics/Publications/PublicationsPolicyAndGuidance/DH_4007697

